# SARS-CoV-2 infections in Italian schools: preliminary findings after one month of school opening during the second wave of the pandemic

**DOI:** 10.1101/2020.10.10.20210328

**Authors:** Danilo Buonsenso, Cristina De Rose, Rossana Moroni, Piero Valentini

**Affiliations:** Department of Woman and Child Health and Public Health, Fondazione Policlinico Universitario A. Gemelli, Rome, Italy; Istituto di Microbiologia, Università Cattolica del Sacro Cuore, Roma, Italia; Global Health Research Institute, Istituto di Igiene, Università Cattolica del Sacro Cuore, Roma, Italia; Direzione Scientifica, Fondazione Policlinico Universitario A. Gemelli IRCCS, Rome, Italy

## Abstract

**Introduction:** The impact of school opening on the SARS-CoV-2 pandemic is still unknown. This study aims to provide preliminary information about the number of SARS-CoV-2 cases among students attending Italian schools.

**Methods:** Data are extracted and analysed from an open access, online dataset that monitor, on a daily basis, media news about SARS-CoV-2 infections of students attending Italian schools

**Results:** As of 5 October 2020, a total of 1350 cases of SARS-CoV-2 infections have been registered in the Italian territory schools (involving 1059 students, 145 teachers and 146 other school members), for a total of 1212 out of 65104 (1.8%) Italian schools involved. National schools reported only 1 case of SARS-CoV-2 infection in more than 90% of cases, and only in one high school a cluster of more than 10 cases have been described (P 0.015). The detection of one or more SARS-CoV-2 infections leaded to the closure of 192 (14.2%) entire schools, more frequently nursery/kindergartens (P<0.0005).

**Discussion:** Our preliminary data support low transmission of SARS-CoV-2 within schools, at least among younger students. However, entire schools are frequently closed in the fear of larger outbreaks. Continuous monitoring of school settings, hopefully through daily updated open access datasets, are needed to better understand the impact of schools on the pandemic, and provide guidelines that better consider different risks within different age groups.

## Introduction

During the first months of the COVID-19 pandemic, starting from April 2020, aiming to reduce the SARS-CoV-2 transmission, most governments declared school closure with approximately 91% of the world’s students in more than 190 countries confined at home (1). This has caused immeasurable disruption to the lives, learning and wellbeing of children around the world. An entire generation has seen its education interrupted (2).

European countries have been able to achieve a good control of the pandemic during summer. Due to the heavy consequences of the restrictive measures on the national economies, restrictive measures have been weakened and most social activities of any type, comprehensively, opened, although masking, hand hygiene and physical distancing was always a necessary obligation according to the governments. However, since September, the number of SARS-CoV-2 cases are progressively increasing in Europe, with countries like France and Spain heavily involved. Italy, the first heavily involved country after China, was one of the countries that best managed the virus during the first wave and successfully lowered cases, particularly during early summer. However, since the first days of September Italy is back to more than 2000 new cases/day with an increase in intensive care unit admissions and deaths. This scenario suggests that the second wave has began in Europe. In this context, “the era of open schools” during the COVID-19 pandemic has started. One of the most important unanswered question is related to school and COVID-19. Will the school reopening have an impact on SARS-CoV-2 transmission among students and the whole community? To date, there are no evidences available to understand the impact of national school opening during the second wave of the pandemic and national governments did not provide yet official data. This information is fundamental to understand how many cases within schools have been notified and if a specific group of students or type of school is more susceptible to SARS-CoV-2 clusters. A better knowledge of SARS-CoV-2 spread among students would improve the school organization during the COVID-19 pandemic, allowing to make decisions that better balance benefits and risk for children and their families.

This study aims to provide preliminary information about the number of SARS-CoV-2 cases among students attending Italian schools, and to assess if the type of school (and, therefore, age groups) are associated with different rates of infection.

## Methods

Data are extracted and analysed from an open access, online dataset that monitor, on a daily basis, media news about SARS-CoV-2 infections of students attending Italian schools (https://bit.ly/covid_scuole; https://www.datawrapper.de/_/DLy9C). See supplementary materials for full details

## Results

As of 5 October 2020, a total of 1350 cases of SARS-CoV-2 infections have been registered in the Italian territory schools (involving 1059 students, 145 teachers and 146 other school members), for a total of 1212 out of 65104 (1.8%) Italian schools involved. The distribution of cases of infections according to school are as follow: 236 (17.5%) pupils from nursery/kindergartens, 300 (22.2%) pupils from elementary schools, 208 (15.4%) pupils from middle school, 452 (33.5%) pupils from high schools and 55 (4.1%) from peer institutions (for 99 children (7.3%) school type was not available).

Students represent the 78.4% of cases. Table 1 shows that the distribution of the categories of infected people is significantly different in the different levels of schools (p <0.0005).

**Table 1.** Distribution of the categories of infected people.

|  |  | Teachers | Students | Others | *Missing* | Total |
| --- | --- | --- | --- | --- | --- | --- |
| Nursery/kindergardens | n | 30 | 171 | 16 | 19 | 236 |
|  | % | 12.7% | 72.5% | 6.8% | 8.1% | 100.0% |
| Elementary | n | 45 | 230 | 7 | 18 | 300 |
|  | % | 15.0% | 76.7% | 2.3% | 6.0% | 100.0% |
| Middle | n | 20 | 177 | 1 | 10 | 208 |
|  | % | 9.6% | 85.1% | 0.5% | 4.8% | 100.0% |
| High school | n | 25 | 384 | 16 | 27 | 452 |
|  | % | 5.5% | 85.0% | 3.5% | 6.0% | 100.0% |
| Peer school | n | 11 | 28 | 7 | 9 | 55 |
|  | % | 20.0% | 50.9% | 12.7% | 16.4% | 100.0% |
| *Missing* | n | 14 | 69 | 5 | 11 | 99 |
|  | % | 14.1% | 69.7% | 5.1% | 11.1% | 100.0% |
| Total | n | 145 | 1059 | 52 | 94 | 1350 |
|  | % | 10.7% | 78.4% | 3.9% | 7.0% | 100.0% |

With the exception of peers schools, National schools reported only 1 case of SARS-CoV-2 infection in more than 90% of cases, and only in one high school a cluster of more than 10 cases have been described (P 0.015, Table 2).

**Table 2.**
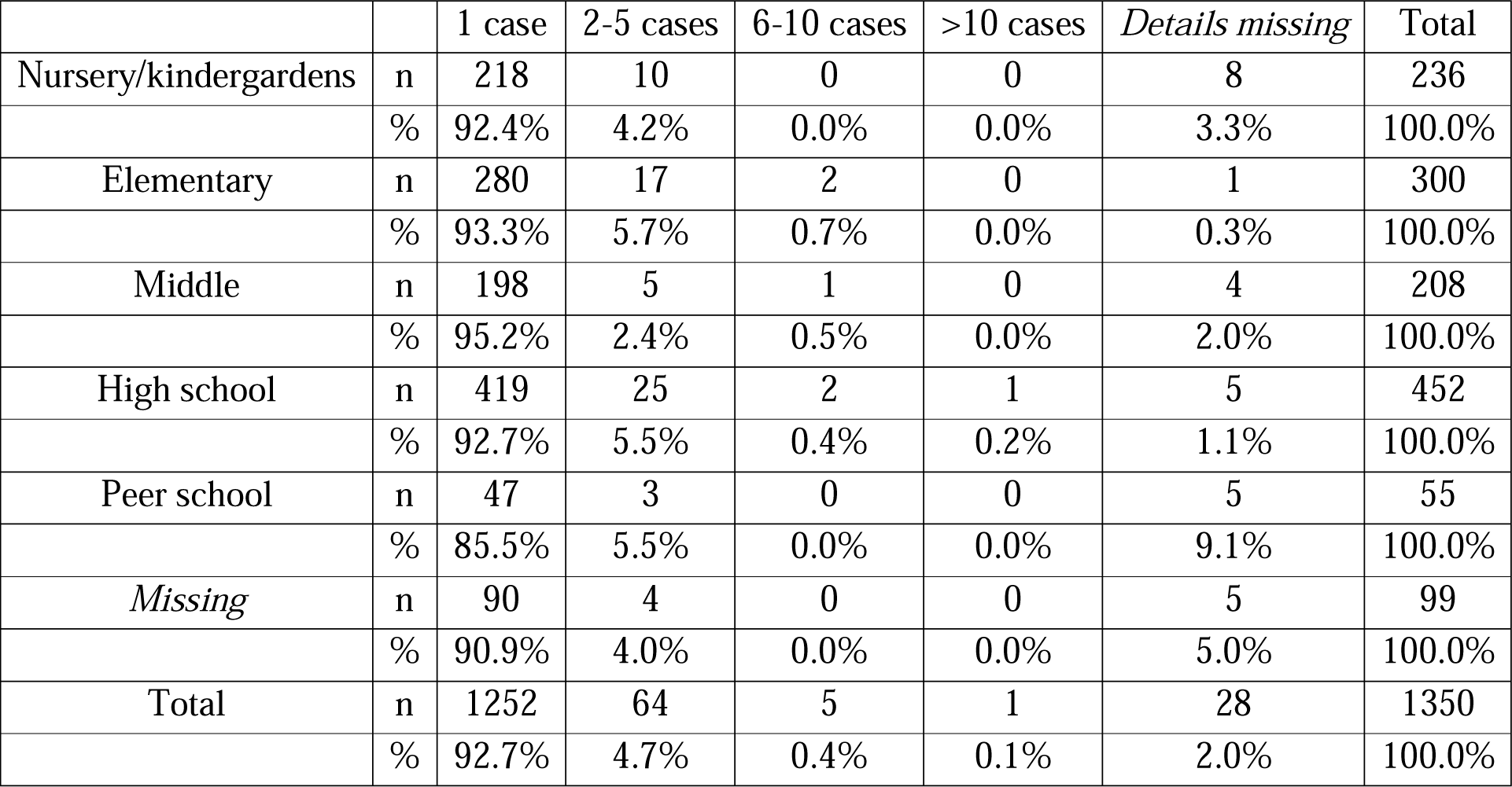

The detection of one or more SARS-CoV-2 infections leaded to the closure of 192 (14.2%) entire schools. In particular, school closures were distributed as follow according to school type (calculated on the total number of each type with at least on infection reported): 51 out of 236 nursery/kindergartens (21.6%); 38 out of 300 elementary schools (12.7%); 23 out of 208 middle schools (11.1%); 49 out of 452 high schools (10.8%); 21 out of 55 peer schools (21.6%) (P<0.0005).

## Discussion

Our preliminary study shows that, as of October 5th, about 1059 students of different age groups have been notified with SARS-CoV-2 infection. Importantly, national schools reported only 1 infection in more than 90% of cases, and only in one high school a cluster of more than 10 cases have been described (P 0.015). These findings would suggest that, when proper preventive measures are respected, the intra-class transmission of the virus is low. This is in agreement with recent studies suggesting that children have lower susceptibility to SARS-CoV-2, compared with adults (4). A systematic review and misanalysis has brought to light stratification of susceptibility of COVID-19 infection of children younger than 14 years old than adults, with adolescents appearing to have similar susceptibility adult. Few data are available on the onward transmission of SARS-CoV-2 from children to others. Data from a large Australian school contact-tracing study (5) suggest that, at a population level, children and adolescents might play only a limited role in the transmission of this virus. This is consistent with the data on susceptibility, suggesting that lower rates of secondary infection mean that children and adolescents have less opportunity for onward transmission. This is consistent with a national South Korean study (6), which found the secondary attack rate from children to household members was extremely low. The available studies suggest children and adolescents play a lesser role in transmission of SARS-CoV-2, which is in marked contrast to influenza (7).

Importantly, despite evidences of a low transmission of SARS-CoV-2 infection among students, and our preliminary findings showing that in most cases only one child has been infected in each school, the detection of one or more infections led to the closure of 192 (14.2%) entire schools. This is in contrast with Italian guidelines (3) and even with the Centers for Diseases and Control operative modalities on the return to school, which suggest quarantine for 14 days for all asymptomatic contacts of a suspicious or positive case (8). School closures in case of small clusters has potential consequences on children and families and, indirectly, on the whole of society. Activating the quarantine for all doubtful or confirmed cases means that we all expect that school continuity is not guaranteed. Not only this has direct consequences on children’s right to education and children’s mental health (2), but also on their families that, having to care of quarantined children, cannot guarantee work continuity. Indirectly, this has obvious consequences on the whole society.

If school closure should not be applied in case of small cluster, does the preventive or routine quarantine for children attending the same class of a child with SARS-CoV-2 infection have any evidence? Already nine months of the pandemic, we have enough evidence that around 1% of all COVID-19 cases involve children (9), that morbidity and mortality are extremely low in this age group (10), and that children play a limited role in viral spread. Children are not COVID-19 super spreaders (11).

Although the data we showed are preliminary, they may confirm that masking, hand hygiene and physical distancing applied in school settings may limit the viral spreading and school or class closures in case of isolated SARS-CoV-2 cases may be a drastic measure that brings low advantages in terms of virus control but fuels an already common sense of precariousness. Rather than seeing at schools as facilitators of the COVID-19 pandemic, we should look at schools as educational settings with a potential of supporting the fight against the pandemic. The fact that children and adolescents are requested to respect certain rules during school hours may trigger positive attitudes even outside school. This is especially important for adolescents which are in need of social relationships but bear a potentially higher susceptibility to SARS-CoV-2 (4).

Considering the heavy indirect consequences of early lockdowns, the social and scientific communities are aware that we must find the proper balance between a near-normal life and the best control of the pandemic. In this balance, we must consider school as well, including children right to education.

Our work has limitations. First, since the dataset is based on news available on the website, it is possible that it underestimates the real situation. Second, the cases in the database are as reliable as the newspapers that publish the news. Although the authors carefully assess the news and collect several information (excluding that news with no enough information), they do not perform a formal investigation to verify the truthfulness or reliability of the news with individual schools. Third, news publishing inevitably suffers from some bias. For example, reports of cases in small towns rather than in large cities seem more frequent, possibly because the former are promptly communicated on the municipal web-pages. Data about contact tracing initiated by the health authorities are missing most times, probably because news agencies prefer to focus on putting more emphasis on situations that see the emergence of an outbreak

In conclusion, our preliminary data suggest that, so far, a limited number of students have been diagnosed with SARS-CoV-2 infection and that intra-class transmission is rare. Active monitoring of the trend and drivers of infections among school is necessary, hopefully with open access data on international platforms, allowing better analyses and comparisons between different settings. Importantly, understanding how different age groups can spread the infection within schools is a priority, since this would allow the development of guidelines more focused on specific school-settings. The goal of our society should be to try to respect, as much as possible, children’s rights.

## Data Availability

all data referred to in the manuscript and note links below are available

## Acknowledgments

We are grateful to Lorenzo Ruffino, university student of the faculty of Economics at the University of Turin, and Vittorio Nicoletta, PhD in Decision-making Systems in Canada for their invaluable work on the database, constantly evolving minute by minute, and for making it in the public knowledge from the very beginning.

