## Supplementary material for "SARS-CoV-2 infections in Italian schools: preliminary findings after one month of school opening during the second wave of the pandemic": Supplemetal Methods

***Study population***

On September 14th, 2020, two PhD students developed an open access, online dataset aiming to monitor, on a daily basis, media news about SARS-CoV-2 infections of students attending Italian schools (<https://bit.ly/covid_scuole>; <https://www.datawrapper.de/_/DLy9C>). At the time of development of this dataset (September 14th, 2020) no similar data were publicly available in Italy. Although the Ministry of Education requested schools to communicate this information on September 25^th^ 2020, at the date of writing of this manuscript (October 9th, 2020) there are no publicly available data about SARS-CoV-2 infections among Italian schools.

The news of cases of SARS-CoV-2 infections are selected from three main resources:

- Google News: the keywords "positive school" are searched. The resulting news is read to select only the relevant ones. For example, the news "School, a positive start" is not considered relevant.

- Institutional sites. Some institutional sites of the Italian Regions provide information on the situation of SARS-CoV-2 infections within schools.

- Reports. Users send news reports through private messages or directly fill the report form. The news reported are read to verify their relevance. If a report is not accompanied by a link which testify the credibility of the website, it is not inserted until a news published by a newspaper or institution is received.

Particular attention is given to news published on the web: national and local newspapers are considered reliable sources, while private blogs are not. All the news is read in its entirety, to extract the variables of interest described below.

The following variables are collected within the dataset: day, region, province, city, school name, type of school, national school code, number of SARS-CoV-2 positive people within the school, category of the positive person (student, teacher, other); type of quarantine activated after a positive case is notified (the whole class, the whole school, students, teachers); school closure (yes, no); number of contacts of the index case resulted positive during epidemiologic assessment; reference of the news.

***The Italian context***

In Italy there are about 65104 schools, including nursery schools, kindergartens, primary schools (elementary), lower secondary schools (middle) and upper secondary schools (high schools), including also the private schools (https://www.tuttitalia.it/scuole). According to the Focus published by the Ministry of Education, with the main data relating to the recently started 2019-2020 school-year, Italian students are now more than 8 million (7599259 students attending National schools, 866,805 attending of peer institutions) (<https://www.miuristruzione.it/13007-quanti-sono-gli-studenti-in-italia-ecco-i-dati-aggiornati-al-2019-2020/>).

The Italian guidelines (3) concerning school opening during the pandemic include: reduced number of students per class or, if this is not feasible, guarantee of a distance of at least 1 meter between each student; frequent hand hygiene; masking. If a child is diagnosed with SARS-CoV-2 infection, the class is quarantined for two weeks, but the rest of the school is requested to continue normal school activities.

***Statistical methods***

Variables present in the database were analysed through descriptive statistical techniques. In particular, the quantitative variables are represented through the following measures: mean and standard deviation. The qualitative variables are represented with absolute frequencies and percentage. The comparison between groups was made with χ2.
